# Treatment of cutaneous leishmaniasis with sodium stibogluconate and allopurinol in a routine setting in Ethiopia: clinical and patient-reported outcomes and operational challenges

**DOI:** 10.1101/2023.07.05.23292210

**Authors:** Saskia van Henten, Fentaw Bialfew, Seid Hassen, Feleke Tilahun, Johan van Griensven, Seid Getahun Abdela

**Affiliations:** Institute of Tropical Medicine, Antwerp, Belgium; Boru Meda Hospital, Dessie, Ethiopia; Wollo University, Dessie, Ethiopia

**Keywords:** *Leishmania aethiopica*, operational research, pentavalent antimonials, treatment extension

## Abstract

Cutaneous leishmaniasis (CL) is common in Ethiopia, but the national guideline does not offer specific treatment recommendations. Consequently, different treatment regimens are used in the country, without quality evidence. In Boru Meda Hospital, sodium stibogluconate (SSG) is routinely used combined with allopurinol for systemic CL treatment, although evidence on effectiveness is limited.

An observational cohort study was done to document clinical treatment outcomes in patients receiving SSG/allopurinol at the end of each 28-day treatment cycle and after 180 days. Patient-reported outcomes were assessed by asking patients to rate lesion severity and by the dermatological life quality index.

A total of 104 patients were included. After one treatment cycle only four patients were clinically cured, although patient-reported outcomes significantly improved. The majority (88) of patients were appointed for a second treatment cycle, of which only 37 (42%) came. Among 36 patients who came for final outcome assessment, 50% was cured. Follow-up and treatment was severely affected by conflict; drug stockouts and insufficient ward capacity for treatment were additional challenges.

Treatment outcomes of SSG/allopurinol were relatively poor, and most patients required more than one cycle of treatment. Shortage of drugs and beds indicates the existing gaps in providing CL treatment in Ethiopia.

## Introduction

Cutaneous leishmaniasis is a common dermatological condition in Ethiopia, where approximately 20,000 – 50,000 patients are affected each year [1]. The causative species is *Leishmania aethiopica,* which typically causes severe lesions on the face that are difficult to treat [2].

Due to lack of good quality evidence for *L. aethiopica* treatment outcomes, the national guideline does not recommend specific treatments [3], but only mentions therapeutic options without specifying the duration. In practice, treatment extension is common [4]–[6], and different combinations of treatment regimens are practiced [4],[6],[7]. As treatment efficacy is species-dependent, there is a dire need to document outcomes of routinely used treatment options for *L. aethiopica*. Intravenous or intramuscular Sodium stibogluconate (SSG) remains the most widely used treatment in Ethiopia. In Boru Meda General Hospital, treatment with systemic SSG is routinely combined with low dose allopurinol, but outcomes have never been reported.

Looking beyond clinician-assessed cure rates, accounting for patient-perspectives is crucial, especially for diseases that negatively affect individuals through psychosocial impact. A recent publication [8] called for including outcomes related to quality of life in studies looking at treatment effectiveness. To date, this has rarely been done around the globe.

In Ethiopia, anecdotal evidence also points to operational challenges such as drug shortages, insufficient bed capacity for inpatient treatment, and poor patient follow-up. Better documentation of these challenges is needed to properly outline and address current gaps in CL treatment. Additionally, parasitological confirmation through demonstration of parasites is required according to the national guideline, while monitoring renal and hepatic function as well as electrocardiography changes is mentioned to evaluate toxicity, without mention of specific frequency. However, it is unclear how often this is done routinely.

In this study, we wanted to document SSG/allopurinol clinical treatment outcomes, determine how often treatment extension was given, and explore how patients reported their outcomes by looking at patient-reported outcomes using the dermatological life quality index, and patient-rated lesion severity. We also described the operational challenges encountered throughout this project.

## Methods

### Setting

The research took place at Boru Meda General Hospital. This hospital which gives specialized dermatology and ophthalmology services with a high caseload of leprosy and CL patients [9]. There is a dermatology ward which has 40 beds. Patients who are confirmed either clinically or microscopically with a skin slit smear are treated either with local treatment by intralesional SSG or cryotherapy (if available) or with systemic treatment.

Routine treatment for severe CL cases that need systemic treatment consists of one or more cycles of 28 days of intramuscular 20mg/kg SSG with a maximum dose of 850mg/day in combination with one 100mg tablet of allopurinol. SSG is given intravenously in case patients cannot tolerate intramuscular injections. Patients are typically admitted for the entire course of treatment. SSG is made available through the World Health Organization via the Ministry of Health.

### Design, population, recruitment, and sample size

This was an observational cohort study among patients with CL who received systemic SSG with allopurinol following routine clinical care decisions. Patients with a clinical or parasitological diagnosis of CL who were started on SSG with allopurinol were invited to the study consecutively. Study recruitment started on 15 February 2021, with the last patient recruited on 04 August 2022, with several interruptions due to political unrest in the area (armed conflict between the Ethiopian government and Tigray militants lasted from 3 November 2020 until 3 November 2022).

Patients visits were planned at baseline (BL) before starting treatment, for each subsequent treatment cycle (Cycle 1 (C1), Cycle 2 (C2) etc.) with study visits planned at the end of each treatment cycle before discharge, and at day 180 (D180) since starting treatment. Patients were also instructed to come back between visits if their lesion did not respond to treatment or was worsening. All information produced during routine diagnosis and treatment was captured, including lab tests that were performed and their respective results.

The original sample size was planned to be 117 patients. Sample size calculation was based on proportion of cure at D180, with a required precision of 10%, using a 95% confidence level, and a conservative estimate of 50% cure. This gave a sample size of 97 patients, which was inflated with 20% to account for loss to follow-up, leading to a final sample size of 117 patients. Due to the conflict and stockouts, recruitment took much longer than planned. Therefore, recruitment was terminated prematurely after a long treatment stockout prevented inclusion of individuals beyond 104 patients.

### Lesion assessment

Before treatment started, the study clinician assessed the lesion by measuring the size of the largest diameter, counting the number of active lesions, classifying the type of CL as localized CL (LCL), muco-cutaneous CL (MCL), and diffuse CL (DCL) [3], and by noting the lesion characteristics (nodular, plaque, ulcerated etc.). The largest lesion was considered as the index lesion.

### Patient-reported outcome measures

The Amharic version of the dermatological life quality index (DLQI) and children’s DLQI (cDLQI) questionnaires [10] (for age between 4-16 years) were administered to patients by the study staff at each study visit (BL, C1, C2 (if applicable), etc., D180). The DLQI questions were not asked to patients below the age of four. For children up to eight years old, the questions were posed to the parent/guardian, while for those aged eight up to twelve years, the questions were asked both to the patient and the parent/guardian. For children above 12, only the child’s answer were considered. Questionnaires were scored and analyzed as recommended [10],[11], with total scores categorized from no effect to extremely large effect and removing questionnaires with answers on eight or less questions as invalid.

Patient-reported outcomes were assessed by asking patients to rate the severity of their lesion on a Likert scale as clear (1), almost clear (2), mild (3), moderate (4) or severe (5) at BL, C1 and any subsequent cycles, and D180.

### Clinical outcome assessment

Lesions were assessed by the clinician at the end of each treatment cycle and at D180. Patients were classified as cured if all lesions that were present at BL had complete reepithelization (if ulcerated) and flattening. Patients were categorized as substantial improvement if not all lesions cured, but there was at least 50% improvement of all lesions compared to BL. Minor improvement required all lesions to have at least 1-49% improvement compared to BL. Worsening was classified compared to the previous lesion assessment, which could be either due to worsening of previously present lesions, or appearance of new lesions. Photographs of the lesions were taken at each study visit to capture the lesion characteristics and outcomes for cross-checking and quality control.

Although we initially planned to include only outcomes 120-240 days after starting treatment, due to the conflict and subsequent travel restrictions, we were more lenient and considered all outcome assessment more than 110 days after starting treatment for D180 outcome.

### Sample collection

This study planned to use the routine skin slit for PCR confirmation, and collected blood and non-invasive tape disc samples (D-Squame, MonaDerm) from each study visit for further analysis.

### Data collection and analysis

A paper data collection form was used to collect patient information which was double entered in a REDCap database [12]. Analysis was done using R version 4.1.3 [13]. To describe the population, numbers, proportions, with medians and interquartile ranges (IQRs) were used. Treatment outcomes were analyzed as categorical variable with multinomial 95% confidence intervals (95% CIs). Cure rate was also analyzed as a proportion with binomial 95% CIs. Differences in DLQI scores and patient global assessment over time were analyzed using the Wilcoxon-signed-rank test. Kappa coefficients were used to measure agreement between when patient’s rated their lesion as clear, and when the clinician rated the lesion as cured.

### Patient tracing

Patient tracing was done over the phone, using patient information (phone number of the patient and at least one family member/neighbor) collected at BL. Patients were called if they were late for an appointment to ask them if they wanted to come. At the end of the study, all patients who missed their D180 visit were called to ask whether they thought their lesion was cured, substantially improved, the same, or worse compared to before and they were asked why they missed their outcome visit.

## Results

A total of 104 patients were included in the study (Supplementary Figure 1), described in Table 1. The majority (85, 81.7%) was referred from health centers or primary hospitals. Patients frequently (74/104, 71.1%) used traditional treatment before coming to the hospital for CL care. Many patients had also tried several modern medications such as ointments and (mostly systemic) antibiotics, but only 6 (5.8%) had previously been treated for CL at a known CL treatment center.

**Table 1.** Baseline characteristics of included patients

|  | **Total N=104** |
| --- | --- |
| Age, median (years) | 18.0 (11.8 - 30.0) |
| Sex, male | 71 (68.3) |
| **CL (Treatment) history** |  |
| Referred from other health facility | 85 (81.7) |
| Previous CL episode (*n*=103) | 6 (5.8) |
| Traditional treatment used | 74 (71.1) |
| Herbal | 64 (61.5) |
| Holy water | 12 (11.5) |
| Burning | 1 (0.9) |
| Other | 3 (2.9) |
| Modern treatment (any) | 40 (38.5) |
| CL-specific treatment | 6 (5.8) |
| **Lesion characteristics** |  |
| Duration in months, median (IQR) | 11.5 (7.0 - 17.0) |
| Type of lesion |  |
| LCL | 30 (28.8) |
| MCL | 69 (66.3) |
| DCL | 5 (4.8) |
| Nr of active lesions |  |
| 1 | 70 (67.3) |
| 2 | 16 (15.4) |
| 3 | 10 (9.6) |
| ≥4 | 8 (7.7) |
| Presence of concomitant CL scar | 5 (4.8) |
| Size (largest diam), median IQR (*n*=100) | 5.0 (4.0 - 8.0) |
| Location index lesion |  |
| Face | 90 (86.5) |
| Arm/hand | 10 (9.6) |
| Other body part | 4 (3.8) |
| Index lesion presentation |  |
| Plaque | 98 (94.2) |
| Scaly | 62 (59.6) |
| Crusted | 55 (52.9) |
| Papular | 54 (51.9) |
| Erythematous | 35 (33.7) |
| Hyperpigmented | 31 (28.9) |
| Swollen | 22 (21.2) |
| Nodular | 21 (20.2) |
| Ulcerated | 18 (17.3) |

*In case of missing data, the total patients with available data are indicated with (n=x) behind the respective variable*

The median duration of symptoms at enrollment into the study was 11.5 months (IQR 7.0 - 17.0). For three quarters (80, 76.9%) the full cost of their admission and treatment was covered by health insurance. Patients came from districts up to 200 kilometer away, although patients more frequently came from areas close to the hospital.

### Patient characteristics

The median age was 18.0 years, with 25% (26) of patients younger than twelve years. More than half of patients (69, 66.3%) presented with mucosal involvement, whereas five (4.8%) were classified as DCL. A severe MCL patient is shown in Figure 1. Most patients had a single lesion, which was most frequently on the face.

**Figure 1.** A patient with severe MCL. A) Swollen, erythematous, and crusted plaques on the face which restricted opening of the eyelid. The patient rated his lesion as severe and had a DLQI score of 16. B) Reduced swelling and crusting with flattening and darkening of the lesions after treatment cycle 1, with substantial improvement as clinical outcome. DLQI score was 9 and patient-reported severity was moderate. C) Further reduction in swelling and plaque-formation with substantial improvement as clinical outcome after cycle 2. DLQI score 8 and patient-assessment mild. D) Lesion was rated as cured with significant remaining scar tissue after cycle 3. Swelling around the eyes has almost completely gone. DLQI was 3 and the patient rated the lesion as mild. Treatment was not extended after the third cycle. E) Worsening with increased swelling, erythema, and papules on both cheeks at D180. The patient rated his DLQI as 20, and his lesion as severe.

### FIGURE OF PATIENT NOT SHOWN IN PREPRINT

### Impact of CL before starting treatment

DLQI data was available for 99 patients before starting treatment, after excluding five invalid questionnaires. Data is shown overall in Figure 2 and separated by the questionnaire used (children when age <15 years and adult for age≥15) in supplementary Table 1. Overall, the median DLQI score was 10 (IQR 5-16). Six (6.1%) patients experienced no effect, 26 (26.3%) a small effect, 22 (22.2%) a moderate effect, 35 (35.4%) a very large effect and ten (10.1%) an extremely large effect. The domain that was most affected both for children (Supplementary Table 2) and adults (Supplementary Table 3) was symptoms and feelings (median score 2.0 (1.0 – 3.0) for children and 3.0 (2.8 – 4.0) for adults). Adults suffered significantly (p<0.001, *Mann Whitney test*) more impact due to CL than children, with a median score of 12.5 (IQR 8.0 - 18.0) compared to 7.0 (IQR 3.0 - 11.0).

**Figure 2.**
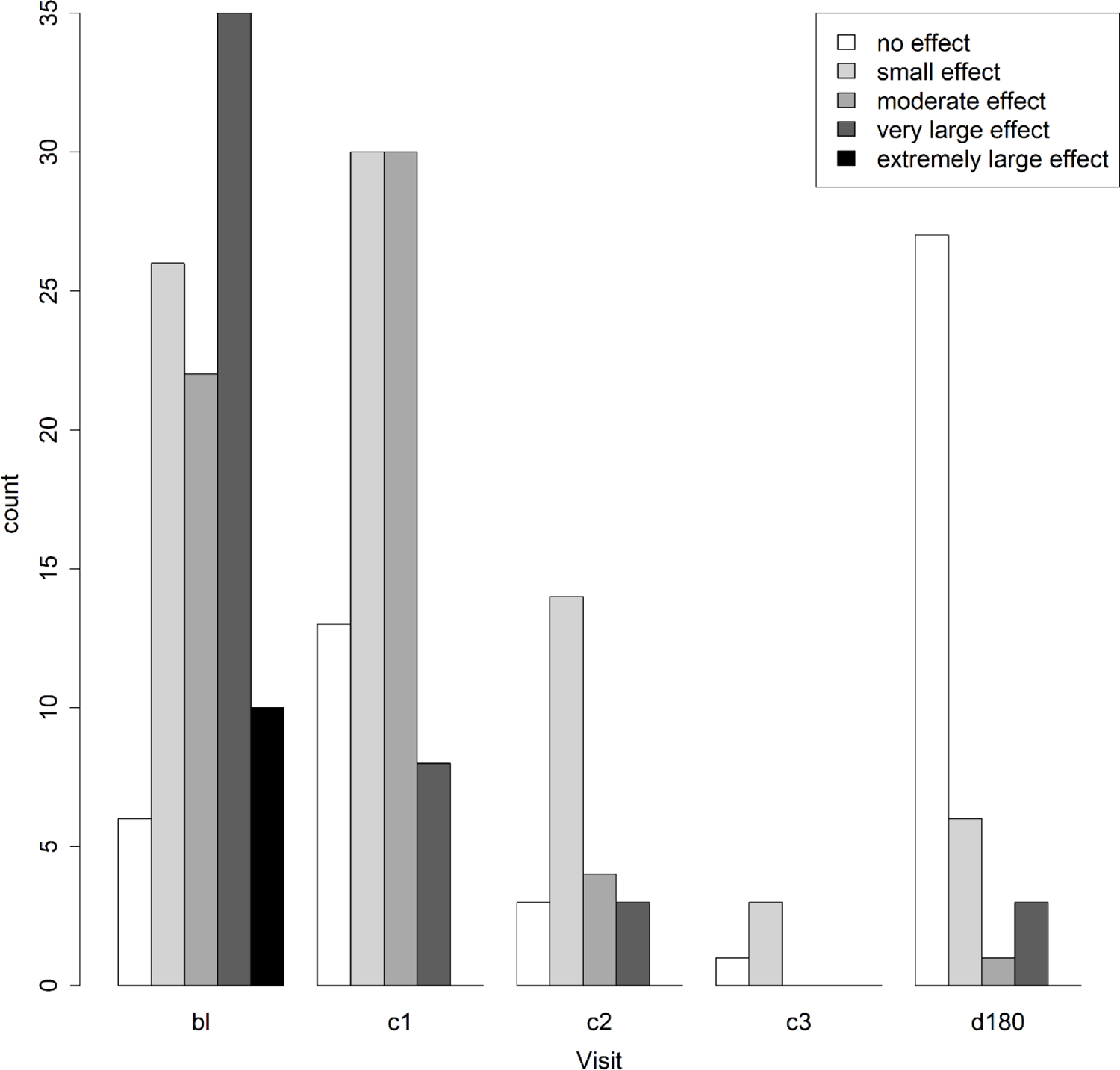
Dermatological life quality index results by study visit. The number of patient in each of the following categories is shown per study visit: no effect, small effect, moderate effect, very large effect, extremely large effect. Results for adults and for children aged 4-15 are combined in this figure. Bl: baseline, c1: after cycle 1, c2: after cycle 2, c3: after cycle 3, d180: after 180 days of starting treatment.

Using patient-reported severity (Table 2), more than half of patients (56, 53.8%) rated their lesions as severe before treatment, with another 38 (36.5%) rating their lesion as moderate.

**Table 2.** Patient-reported severity assessment at the different study visits

| Category | Baseline Total (%)<br>N=104 | End of Cycle 1 (%)<br>N=89 | End of Cycle 2 (%)<br>N=24 | Day 180<br>N=32 (30.8) |
| --- | --- | --- | --- | --- |
| Clear | 0 (0) | 1 (1.1) | 3 (12.5) | 11 (34.4) |
| Almost clear | 0 (0) | 18 (20.7) | 7 (29.2) | 8 (25.0) |
| Mild | 9 (8.7) | 39 (44.8) | 10 (41.7) | 5 (15.6) |
| Moderate | 38 (36.5) | 28 (32.2) | 4 (16.7) | 4 (12.5) |
| Severe | 56 (53.8) | 1 (1.1) | 0 (0) | 4 (12.5) |
| *Missing<sup>a</sup>* | *1/104 (1.0)* | *2/89 (2.2)* | *1/24<sup>b</sup> (4.2)* | *4 (11.1)* |
| Median | 5.0 (4.0 - 5.0) | 3.0 (3.0 - 4.0) | 3.0 (2.0 - 3.0) | 2.0 (1.0 – 3.3) |

<sup>a</sup>Missing proportions are calculated taking the total number of patients who came for that visit
<sup>b</sup>Patient global assessment was taken from one patient for whom the clinical assessment was missed, but missed from one patient who had a clinical outcome

### Laboratory testing

A skin slit smear test was done to confirm CL in almost all (100, 96.2%) patients but it was negative in 56 (56.0%) patients. Although two additional patients were confirmed by fine needle aspiration cytology, the majority of patients (58, 55.8%) was treated based on clinical diagnosis alone. Although complete blood count (CBC) and organ function tests were routinely requested before starting treatment, they were done for only 39 (37.5%) and 24 (23.1%) of patients at the start of the first treatment cycle. HIV tests were done for a total of 22 patients (21.2%), and for only one of the five DCL patients; all patients tested were HIV negative.

During the first cycle of treatment, CBC was done for half of the patients (53, 51.0%). Six (5.8%) patients had more than one CBC result. Blood count was mostly tested in the middle of the treatment period. Organ function tests was done in C1 for less than a quarter of patients (24, 23.1%). ECG was done for only two patients.

### Treatment and follow-up

A total of 99 patients completed their first treatment cycle of SSG/allopurinol combination treatment, of which 89 completed all C1 study procedures (Supplementary Figure 1). Most missed C1 study visits took place during fall 2020, when the hospital was overloaded with soldiers from the front, and study staff were unable to perform their study activities next to their routine engagements. Among 88 patients appointed for retreatment at C2, only 37 (42.0%) actually came for a second treatment cycle during the study period (Supplementary Figure 1). One patient suddenly died in the middle of her second cycle, which could be potentially related to cardiotoxicity of SSG, but since no ECG was done this cannot be confirmed. Three patients were discharged before finishing their C2 since the hospital was evacuated. For nine study patients, procedures for the C2 study visit were missed, mainly due to the conflict. Only around one third (36/104, 34.6%) of patients came for their D180 outcome assessment.

### Clinical treatment outcomes

Clinical outcomes are shown in Table 3. Among the 89 patients that completed C1 study procedures, 4 (4.5%) were cured after 28 days, 71 (79.8%) had substantial improvement, 12 (13.5%) minor improvement, 1 (1.2%) had no improvement, and another 1 (1.2%) worsening. The majority 83/89 (92.8%) of patients were appointed for another cycle of treatment (Supplementary Figure 1).

**Table 3:** Clinical treatment outcomes at the different study visits

| Category | C1 (%) |  | C2 (%) |  | C3 (%) | D180 (%) |  |
| --- | --- | --- | --- | --- | --- | --- | --- |
|  | N=89 (85.6) | 95% CI | N=24 (23.1) | 95% CI | N=4 (3.8) | N=36 (34.6) | 95% CI |
| Cure | 4 (4.5) | 0 – 13.1 | 2 (8.3) | 0 – 24.7 | 2 (50.0) | 18 (50.0) | 36.1 – 68.4 |
| Substantial improvement | 71 (79.8) | 73.0 – 88.3 | 20 (83.3) | 75.0 – 99.7 | 2 (50.0) | 11 (30.6) | 16.7 – 48.9 |
| Minor improvement | 12 (13.5) | 6.7 – 22.1 | 2 (8.3) | 0 – 24.7 |  | 2 (5.6) | 0 – 23.9 |
| No improvement | 1 (1.1) | 0 – 9.7 | 0 (0) | 0 – 16.4 |  | 0 (0) | 0 – 18.3 |
| Worsening | 1 (1.1) | 0 – 9.7 | 0 (0) | 0 – 16.4 |  | 5 (13.9) | 0 – 32.3 |
| Missing<sup>a</sup> | 10 (10.1) |  | 9 (27.3) |  | 0 | 0 |  |

<sup>a</sup>Missing are calculated from the total number of patients who completed the treatment cycle. Which was 99 for C1, 33 for C2, and 4 for C3.

After finishing the second treatment cycle, the majority of patients with a known outcome (20/24, 83.3%) had substantial improvement, 2 (8.7%) were cured, and 2 (8.7%) had minor improvement (Table 3). For the majority of treated patients (21/24, 87.5%), another treatment cycle was deemed necessary (Supplementary Figure 1).

Half of the patients who came for their D180 visit (18/36, 50.0%) were cured, 11 (30.6%) had substantial improvement, 2 (5.6%) minor improvement and 5 (13.9%) worsening. Four study patients were continued on treatment after finishing their D180 visit. Outcomes were significantly better (p=0.032) in the group who came according to the ordered treatment extension, with eight of ten with a D180 outcome cured, while this was ten out of 25 (40.0%) in those who did not come for planned treatment extension. Still, these ten patients who cured, and another ten who had substantial improvement, indicating that further self-healing of the lesion can still take place even if further treatment is deemed necessary.

### Patient-reported outcome data

Patient-reported outcomes are shown in Table 2. After one cycle, patients’ rated their lesion severity as significantly improved compared to BL (p<0.001, *Wilcoxon-Signed-Rank test*), with a median score of 3.0 (IQR 3.0 – 4.0) compared to 5.0 (IQR 4.0 – 5.0) at BL.

At D180, around one third (11/32, 34.4%) of patients reported their lesions to be cleared, one quarter (8/32, 25.0%) said they were almost cleared, five (15.6%) said they were mild, four (12.5) assessed them to be moderate and four (12.5) as severe. The median score given at D180 was significantly lower than at BL (p<0.001, *Wilcoxon-Signed-Rank test*).

Overall, the proportion of patients cured according to the clinician was significantly different from the patient-reported clearance rate (Table 4). Clinician cure rate was higher (13.0 vs 8.3) at C2, but lower than patient-reported clearance rate at C1 (4.5 vs 1.1) and D180 (50.0 vs 34.4) respectively. Agreement measured by Kappa’s coefficient between clinical cure and patient-reported cure was 0.69 at D180 and 0.78 at C2. At C1, this was much lower, at 0.38.

**Table 4.**
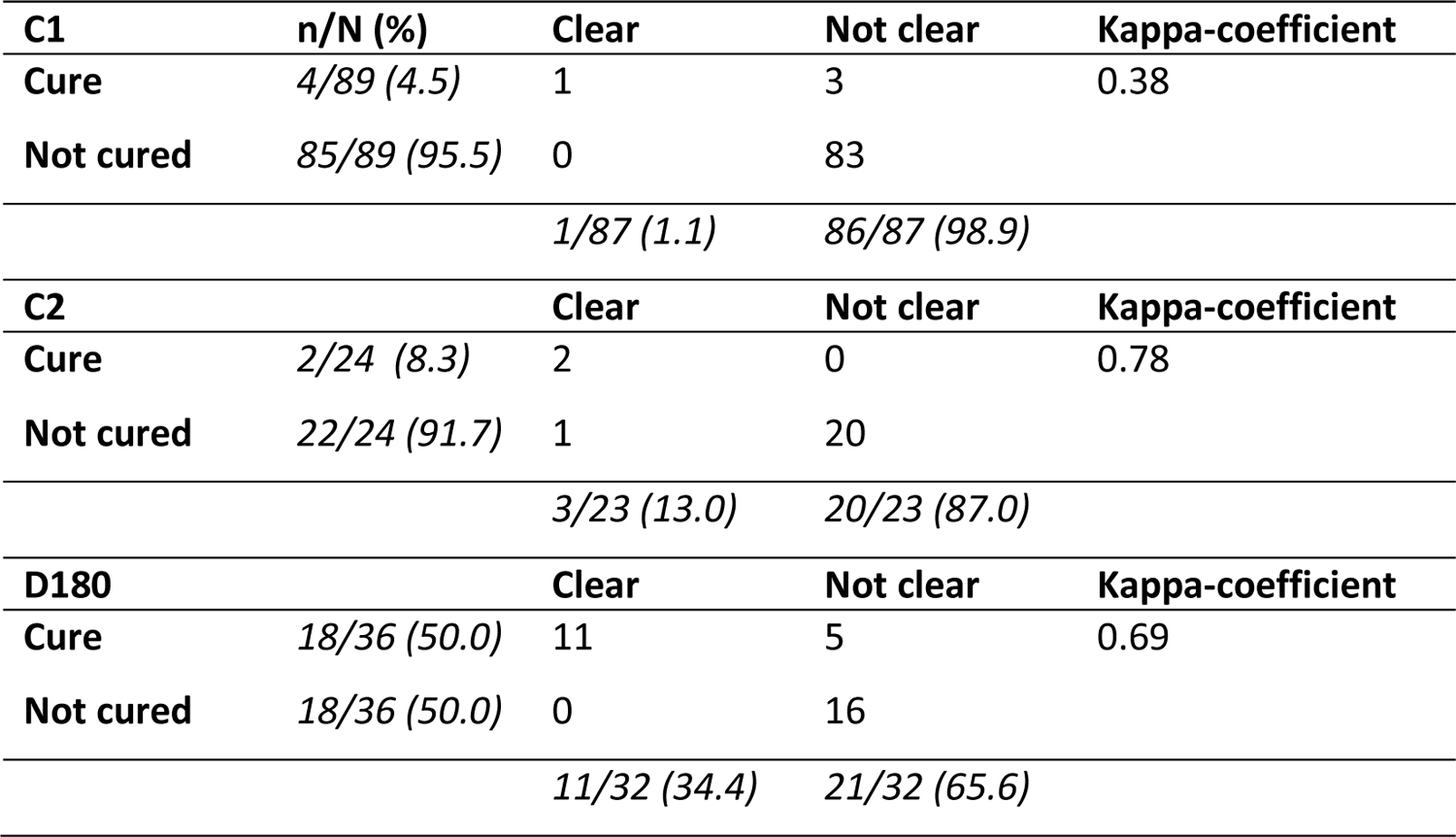
Agreement between clinical and patient-reported outcomes

| **C1** | **n/N (%)** | **Clear** | **Not clear** | **Kappa-coefficient** |
| --- | --- | --- | --- | --- |
| **Cure** | 4/89 (4.5) | 1 | 3 | 0.38 |
| **Not cured** | 85/89 (95.5) | 0 | 83 |  |
|  |  | 1/87 (1.1) | 86/87 (98.9) |  |
| **C2** |  | **Clear** | **Not clear** | **Kappa-coefficient** |
| **Cure** | 2/24 (8.3) | 2 | 0 | 0.78 |
| **Not cured** | 22/24 (91.7) | 1 | 20 |  |
|  |  | 3/23 (13.0) | 20/23 (87.0) |  |
| **D180** |  | **Clear** | **Not clear** | **Kappa-coefficient** |
| **Cure** | 18/36 (50.0) | 11 | 5 | 0.69 |
| **Not cured** | 18/36 (50.0) | 0 | 16 |  |
|  |  | 11/32 (34.4) | 21/32 (65.6) |  |

### Dermatological life quality index after treatment

Results from the DLQI data are shown in Figure 2. Baseline and C1 data was available for 79 patients, whose median score was 9 (IQR 5-15) at BL and 6 (IQR 3-8) at C1. After treatment, the DLQI scores of the 79 patients with BL and C1 data decreased significantly (p<0.001, *Wilcoxon-Signed-Rank-test)*, with a median score of 6.0 (IQR 3.0 - 8.0) at the C1 study visit, compared to 9 (IQR 5-15) at BL. At D180, the median DLQI score of patients was 0 (IQR 0-2), with 27 (73.0%) patients having no effect, six (16.2%) a small effect, one (2.7%) a moderate effect, and three (8.1%) a very large effect.

### Tracing patient outcomes

Because so few patients came for the outcome assessment visit, we systematically called up all patients to ask how they perceived their lesion now, and why they did not come for their D180 visit.

We were able to reach three quarters (52/67, 77.6%) of the missed patients by phone. Over half (29/52, 55.8%) of them reported that their lesion was cured, around a third (17/52, 32.7%) said there was improvement, and four patients (7.7%) reported no change or worsening. The most common reasons the D180 visit was missed was because of the conflict (13/52, 25%), because the lesion was deemed to be cured already (12/52, 23.1%), and due to lack of money or time (10/52, 19.2%). Three patients reported they did not come because further treatment was not available.

### Operational challenges

Many operational challenges were encountered during the study. Drug stockouts occurred three times: allopurinol was unavailable for about one month in April 2021, SSG stockouts occurred twice, once for six weeks from late March-2022 until 8 May 2022, and for a second time from the 12th of august 2022 until the end of the study (over 3.5 months), which also led to early termination of recruitment.

Due to the conflict between the Tigray militants and the national armed forces, the hospital became one of the main treatment sites for wounded soldiers. As the front advanced further south, the hospital staff was forced to evacuate on the 14th of October 2021. During the occupation which lasted several months, the study freezer including all study samples and camera were stolen, and several documents were lost. Hospital services were restarted from February 2022 onwards.

Another challenge was that the patients requiring treatment exceeded the ward capacity. In October 2022 there was a list of 171 patients waiting for admission for CL treatment. Of these, 24 were study patients.

## Discussion

We report on the routine use of systemic SSG in combination with allopurinol in Ethiopia. At the end of the first treatment cycle, only four patients were considered cured, whereas almost all other patients were appointed for another treatment cycle. Less than half of appointed patients returned for further treatment, and only a small number of patients came for their final outcome assessment visit, where around half of them were cured and around a third had substantial improvement. According to their own assessment, one third said their lesion had cleared, but around 70% said their lesion no longer had an impact on their quality of life. In addition, this study highlights the effects of the conflict in Northern Ethiopia on treatment provision for CL, but also shows that drug stockouts and shortage of beds - problems not directly related to the conflict-impact treatment.

Most patients had insurance [14] which covered the cost of treatment, and since patients need to be referred in order to qualify for health insurance to cover hospital costs, the majority was referred from other health facilities. However, the fact that most patients still came with longstanding lesions after trying traditional treatment and various other modern treatments, highlights delays in seeking care. This likely contributes to larger lesion size and more severe scar formation, thereby aggravating the psychosocial impact of CL.

In this patient population getting systemic treatment, the impact of CL on their quality of life prior to treatment was high, with 35% of patients indicating a very large, and 10% an extremely large impact. More than half of the patients rated their lesion as severe before treatment. This clearly shows the big impact CL has on these patients, and emphasizes the need for treatment and psychosocial support. Although clinical cure rates at the final visit were relatively poor around 50%, an additional 30% had substantial improvement and patient-reported outcomes at this time indicated a significant improvement in their quality of life as well as their severity rating of their lesion. Although social desirability bias may play a role, our findings seem to highlight that even though the proportion of patients cured is low, most patients experience a significant improvement in their lesion appearance and associated quality of life. Therefore, treatment still seems to have a positive impact on patients.

In this setting, systemic SSG 20mg per kg with a maximum of 850mg/day is routinely used with low dose allopurinol of 100mg per day, without a clear evidence-base. Evidence on the use of allopurinol for treatment of CL is generally conflicting. Clinical trials in Iran [15] and Colombia [16],[17] showed that allopurinol significantly increased the effectiveness of antimonials, and that this combination can cure lesions that failed to respond to other treatments [18]–[20]. On the other hand, randomized clinical trials in Peru [21] and Iran [22] showed no added benefit of allopurinol compared to antimonials alone. However, almost all previous studies used allopurinol at a high dose of 20mg/kg, adding up to a daily dose usually exceeding 1000mg-in stark contrast with our study where patients received a daily allopurinol dose of only 100mg. This low-dose was started based on expert opinion without proper evidence, and has been continued since only at this center, further underlining the need for clear evidence-based treatment guidelines.

Due to the low number of patients with final treatment outcomes, we should be careful to draw conclusions about effectiveness of SSG/low-dose allopurinol. Poor follow-up data is partially caused by the conflict, but also related to treatment outcomes, as both patients with very poor outcomes and those already cured are unlikely to come to follow-up. However, informal outcome data collected by calling patients indicates that in patients who missed their outcome visit, the proportions cured and improved were similar to those who attended their outcome visit. Cure rates are similar to what our group reported for 28 days of miltefosine treatment [23]. Since the majority of the cured patients in this study were hospitalized for at least two treatment cycles of SSG with allopurinol, miltefosine seems preferable, as patients took their treatment at home after one week of hospitalization.

The decision to extend treatment was very common, although many patients never actually returned for further treatment. It is hard to draw conclusions on the outcomes of treatment extension, as the lesion severity and response to the first treatment cycle is likely related to patients coming for follow-up. Since more than half of the patients who did not come for their appointed treatment extension were still cured at D180, treatment extension does not always seem needed. However, at Boru Meda hospital, due to fear of relapse, treatment is typically extended if the lesion is not cured, which is assessed directly or two weeks after completing the first treatment cycle. Some argue that this is too early, as tissue repair is thought to still take several weeks after parasites have been killed [24]. Formal randomized studies are needed to assess the additive effect of treatment extension, but consensus on when and for whom treatment extension is needed could also provide more clarity.

Our study highlights several important operational challenges of treating CL in Ethiopia. Drug stockouts were frequently seen, indicating that drug supplies are not consistently reliable. SSG is mainly supplied by WHO through the Ministry of Health, but it seems the supply does not match the current needs of CL treatment centers. Although anti-leishmanial drugs are listed in the essential drug list of the country[25], consistent supply should be ensured. The impact of war on CL treatment was large, as hospital services were shut down for months, treatments were interrupted, and follow-up was perturbed. Unavailability of drugs and interruption of services probably contributed to the long waiting list for inpatient treatment. Insufficient treatment capacity further interferes with treatment schedules and follow-up, as most patients who were reappointed for treatment did not come, or could not be treated due to lack of space. Ambulatory treatment with oral miltefosine as an oral drug could solve part of the problem, although cost and access are issues that currently limit widespread implementation.

A recent study in Ethiopia highlighted that there is severe underreporting of CL, and that the majority of patients never seek care at modern treatment facilities [26]. According to the 2030 Neglected Tropical Disease (NDT) roadmap [27], 85% of all CL cases should be detected and 95% of them should be treated, which would increase patient flow to treatment centers. The long waitlist for inpatient treatment indicates treatment facilities may not have the capacity needed to treat the CL caseload and meet the roadmap targets. To succeed, there should either be a great scale-up of referral treatment sites if inpatient treatment with systemic antimonials be continued. Boru Meda has recently upgraded their dermatological treatment capacity with 16 beds, but waiting lists for treatment persist. Rather, innovative approaches such as decentralization should be explored to meet increasing demands of CL diagnosis and care. These could follow the multi-drug resistant tuberculosis system, where patients are diagnosed at the referral center, but take treatment at lower level health facilities closed to their home [28].

Although the guidelines advise parasitological confirmation of patients before treatment, in practice many patients are treated empirically. Data from a previous study in a similar population showed that this approach seems sound, as 34/40 (85%) of microscopy negative patients were confirmed by PCR [23]. Unfortunately, we could not confirm the diagnosis by PCR in this study as samples were lost during the hospital’s occupation. Other lab tests were not routinely performed, and ECG was rarely done. There was one patient who died during the second treatment cycle. Since SSG is known to cause QTc interval prolongation, [29], we cannot exclude the death was due to SSG-related QTc prolongation or arrhythmias which could have been identified on ECG. This case emphasizes the need of ECG monitoring at least before the start of treatment. Cheap and portable six-lead ECG devices attached to mobile phones were recently FDA approved for measuring QTc intervals [30], and could facilitate clearance for treatment with SSG at the health center or primary hospital.

This study had several limitations. Since photographs were lost due to the stolen camera, it was impossible to systematically obtain a second opinion for outcome assessment of patients. Therefore, outcome assessment is solely based on the opinion of the treating clinician, which is subjective, although this reflects what is routinely done. In future studies, we suggest outcome assessment be done by two independent dermatologists, preferably in person so flattening can be addressed properly by palpation. Second, final outcome data could be obtained for only a small number of patients, which affects the validity of our outcome data. Although lost-to-follow-up for final outcomes was very high, most patients could be reached by phone. As patient-reported outcomes were correlated with clinical outcomes, this seems an opportunity to supplement outcome data for future studies investigating treatment effectiveness.

## Conclusions

Treatment with SSG and low-dose allopurinol led to low cure rates after one cycle, with frequent treatment extension. After 180 days, around 50% of patients had clinical cure and 30% improvement, while patient-reported outcomes significantly improved. Treatment stockouts were frequent, and bed capacity was limited leading to long waiting lists for treatment. Increasing treatment capacity or decentralization to lower level health facilities seem vital to meet the WHO NTD roadmap goals aimed to improve access to diagnosis and treatment for cutaneous leishmaniasis in Ethiopia.

## Author contributions

Conceptualization: SvH, JvG, SGA; Methodology, SvH, JvG, SGA; Formal Analysis: SvH; Investigation: FB, SH, FT; Resources: JvG; Data Curation: FB, SH, FT; Writing – Original Draft Preparation: SvH; Writing – Review & Editing: JvG, SGA; Visualization: SvH; Supervision: SvH, SGA; Project Administration: SvH, SGA, FT; Funding Acquisition, JvG.

## Institutional Review Board Statement

The study was conducted according to the guidelines of the Declaration of Helsinki, and approved by the ethical review committees of the Institute of Tropical Medicine, Antwerp (Ref 1351/20, 23 March 2020); the University hospital of Antwerp (Ref 20/15/183, 27 April 2020), and Wollo University College of Medicine and Health Sciences Ethics committee, Dessie (Ref 127/02/13, 14 October 2020).

## Informed consent statement

Informed consent was obtained from subjects involved in the study, or from the guardian if patients were below 12 years old. Assent was additionally obtained from children aged 12-17. Patients specifically consented to having their photograph taken and for it to be shared - granted they could not be identified -, and for patient tracing by phone. The study was registered at ClinicalTrials.gov as NCT04699383.

## Data Availability

Data will not be made openly accessible due to ethical and privacy concerns. Data can however be made available after approval of a motivated and written request to ITMs Research Data Access Committee.

## Acknowledgements

We want to thank the patients who agreed to participate in this study, and all the Boru Meda staff who were involved in data collection and processing. We would also like to thank University of Gondar staff Mezgebu Silamsaw and Tarekegn Asmamaw and ITM staff Myrthe Pareyn and Gudrun van Belle who helped with project administration and financing.

