## Supplementary materials for "Treatment of cutaneous leishmaniasis with sodium stibogluconate and allopurinol in a routine setting in Ethiopia: clinical and patient-reported outcomes and operational challenges"

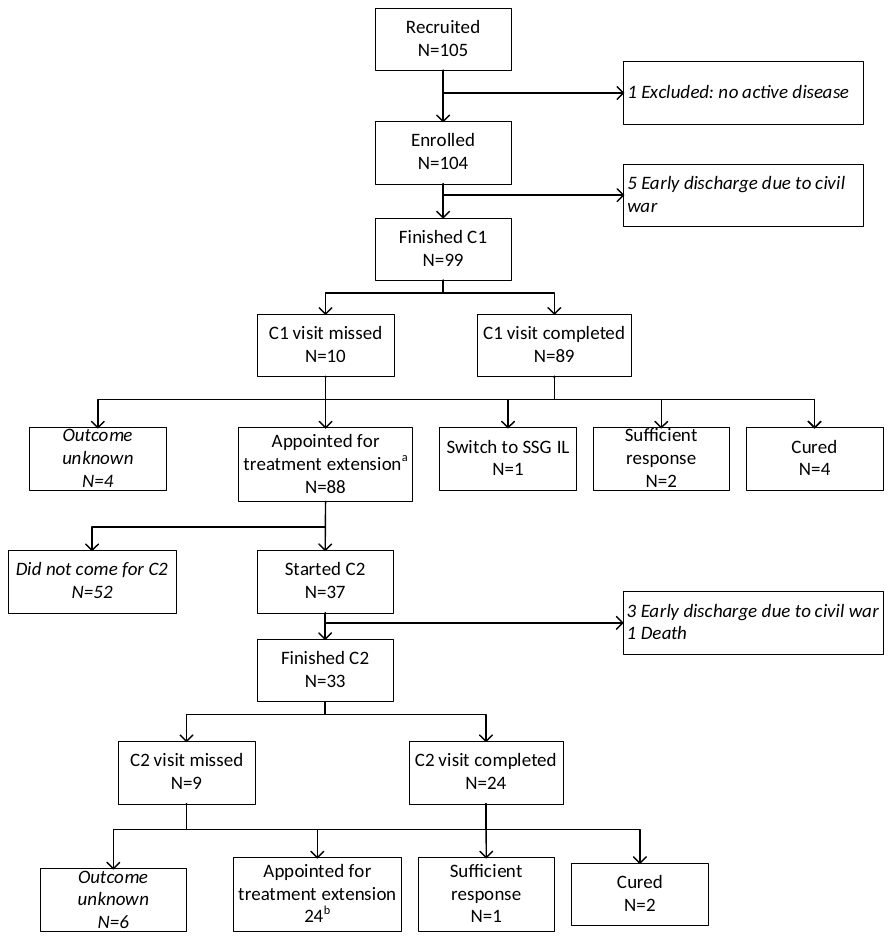

**Supplementary Figure 1.**

**^a^**Among those extended, 82 completed C1 study visits and 6 did not. ^b^Among those extended after C2, 21 completed C2 study visits and 3 did not.

| **Category** | **Total (%)**  **N=99** | **Adults**  **N=56** | **Children**  **N=43** |  |
| --- | --- | --- | --- | --- |
| No effect | 6 (6.1) | 0 (0) | 6 (14.0) |  |
| Small effect | 26 (26.3) | 11 (19.6) | 15 (34.9) |  |
| Moderate effect | 22(22.2) | 9 (16.1) | 13 (30.2) |  |
| Very large effect | 35 (35.4) | 30 (53.6) | 5 (11.6) |  |
| Extremely large effect | 10 (10.1) | 6 (10.7) | 4 (9.3) |  |
| *Invalid* | *4 (3.8)* |  |  |  |
| Median | **10.0 (5.0 - 16.0)** | 12.5 (8.0 – 18.0) | 7.0 (3.0 – 11.0) | <0.001 |
| Mean | **10.7** |  | 7.8 |  |

**Supplementary Table 1: Impact of cutaneous leishmaniasis on the dermatological life quality index before treatment for adults and children separately**

| **Category** | **No effect (0-1)** | **2-3 points** | **4-6 points** | **Median score** |
| --- | --- | --- | --- | --- |
| Symptoms and feelings (0-6) | 5 (8.9) | 27 (48.2) | 24 (42.9) | 3.0 (2.8 - 4.0) |
| Daily activities (0-6) | 15 (26.8) | 26 (46.4) | 15 (26.8) | 2.0 (1.0 -4.0) |
| Leisure (0-6) | 21 (37.5) | 25 (44.6) | 10 (17.9) | 2.0 (1.0 - 3.0) |
| School/work (0-3) | 24 (42.9) | 32 (57.1) | - | 2.0 (0 - 3.0) |
| Personal relationships (0-6) | 26 (46.4) | 16 (28.6) | 14 (25.0) | 2.0 (0 - 3.2) |

**Supplementary Table 2. Quality of life domains affected for adults at baseline (n=56)**

**Supplementary Table 3. Quality of life domains affected for children at baseline (n=43)**

| **Category** | **No effect (0-1)** | **2-3 points** | **4-9 points** | **Median score** |
| --- | --- | --- | --- | --- |
| Symptoms and feelings (0-6) | 21 (48.8) | 15 (34.9) | 7 (16.3) | 2.0 (1.0 - 3.0) |
| Leisure (0-9) | 25 (58.1) | 6 (14.0) | 12 (27.9) | 1.0 (0 - 4.0) |
| School/work (0-3) | 28 (65.1) | 15 (37.2) | - | 1.0 (0 - 3.0) |
| Personal relationships (0-6) | 30 (69.8) | 12 (27.9) | 1 (2.3) | 0 (0 - 2.0) |
| Sleep (0-3) | 38 (88.4) | 5 (11.6) | - | 0 (0 - 0) |
